# Deep multimodal clustering identifies biological subtypes of normal aging with divergent cognitive decline risk

**DOI:** 10.64898/2026.08.18.26360683

**Authors:** Monica M. Diaz, Eran Dayan, The Health and Aging Brain Study (HABS-HD) Study Team, The Alzheimer’s Disease Neuroimaging Initiative

## Abstract

Cognitively normal older adults are often regarded as a homogeneous population in preventive and disease-modifying clinical trials for dementia. However, longer-term cognitive aging outcomes vary substantially in this population, and this variability remains poorly understood. Here, we leveraged rich multimodal, multi-domain biomarker profiles from a large prospective cohort (N=1,136), and Deep Embedded Clustering, to cluster cognitively normal older adults into biologically distinct subgroups. Input data included cortical thickness derived from MRI, plasma Alzheimer’s disease (AD) biomarkers, plasma inflammatory biomarkers, and vascular measures. The deep clustering algorithm identified three biologically distinct subgroups within the sample, stratified along a gradient of neurobiological burden (low, intermediate, and high). Cortical thinning and inflammatory burden were the primary drivers of clustering assignments. The High-Burden subgroup showed significantly worse memory and executive function, elevated cardiometabolic comorbidity, and markedly higher rates of conversion to mild cognitive impairment or dementia within two years. The results were validated in an independent external sample. The study reveals marked variability among individuals who are otherwise all defined as cognitively normal, and provides a data-driven stratification framework for enriching disease-modifying and preventive trials by identifying cognitively normal individuals at high risk for future cognitive decline.

## Introduction

Healthy aging varies across individuals, with large inter-individual variability observed in cognitive aging trajectories^1,2^. While some individuals maintain preserved cognition over time, others demonstrate more accelerated cognitive aging and are often at higher risk for cognitive decline and dementia^3,4^. High risk for cognitive decline and dementia can be predicted from multiple risk factors^5^, and may feasibly reflect complex interactions among multiple neurobiological mechanisms^6,7^. However, it remains poorly understood whether the co-occurrence and interactions of neurobiological risk mechanisms can meaningfully differentiate cognitively normal individuals, and predict cognitive decline and dementia.

Among many of the mechanisms studied in the literature, cortical thinning, vascular comorbidities, Alzheimer’s disease (AD)-related proteopathic burden and elevated inflammation have been shown to particularly impact brain aging trajectories^8–10^. Namely, variation in regional measures of cortical thickness can be found in individuals with normal cognition^11^ and is predictive of cognitive decline^12^. Vascular contributions to cognitive decline and dementia are very well documented^13,14^, with significant variation observed in the extent to which vascular burden impacts cognitive outcomes in older adults^15^. Proteopathic burden is another widely studied contributor to cognitive aging outcomes and dementia risk. Studies have identified significant heterogeneity in AD biomarkers among individuals with normal cognition^16^, and levels of plasma biomarkers for amyloid and tau pathology are predictive of cognitive decline in this population^17–19^. Finally, microglial activation and elevated inflammatory signaling have been linked to cognitive decline and other age-associated changes, such as cortical atrophy and white-matter alterations^20,21^. Elevated inflammatory markers have also been associated with neurodegenerative changes in individuals without clinically detectable neurodegeneration^22^, suggesting that inflammation may modulate how biological aging processes translate into clinically detectable cognitive impairment.

Despite evidence that aging trajectories are impacted by multiple interacting biological pathways, such as those detailed above, most studies to date have focused on single putative mechanisms, which may not account for the heterogeneity of factors that impact cognitive trajectories as people age. Advances in machine/deep learning methods allow for the identification of subtypes using high-dimensional data, yet the majority of studies utilizing these and other data-driven methods have focused on subtyping and stratification of AD or mild cognitive impairment (MCI)^23–27^, leaving cognitively normal aging largely understudied.

To identify meaningful subgroups within cognitively normal aging, while addressing the inherent high dimensionality and heterogeneity of multimodal data from distinct neurobiological domains, we applied a modified Deep Embedded Clustering (DEC)^28^ (**Fig. 1**). In this approach, a deep autoencoder is applied to learn a compressed non-linear latent representation of the multimodal set of features, and these latent representations are simultaneously used to optimally cluster subjects into subgroups. Phenotypic data, unseen by the DEC model, were then used to validate the clustering-based subgroups, and the results were further validated against an external dataset. We hypothesized that multimodal clustering would reveal biologically distinct cognitive risk groups and that clusters characterized by cortical thinning, increased proteopathic burden, elevated inflammation and increased vascular burden would be associated with worse cognitive performance and greater risk of subsequent cognitive decline.

**Figure 1.**
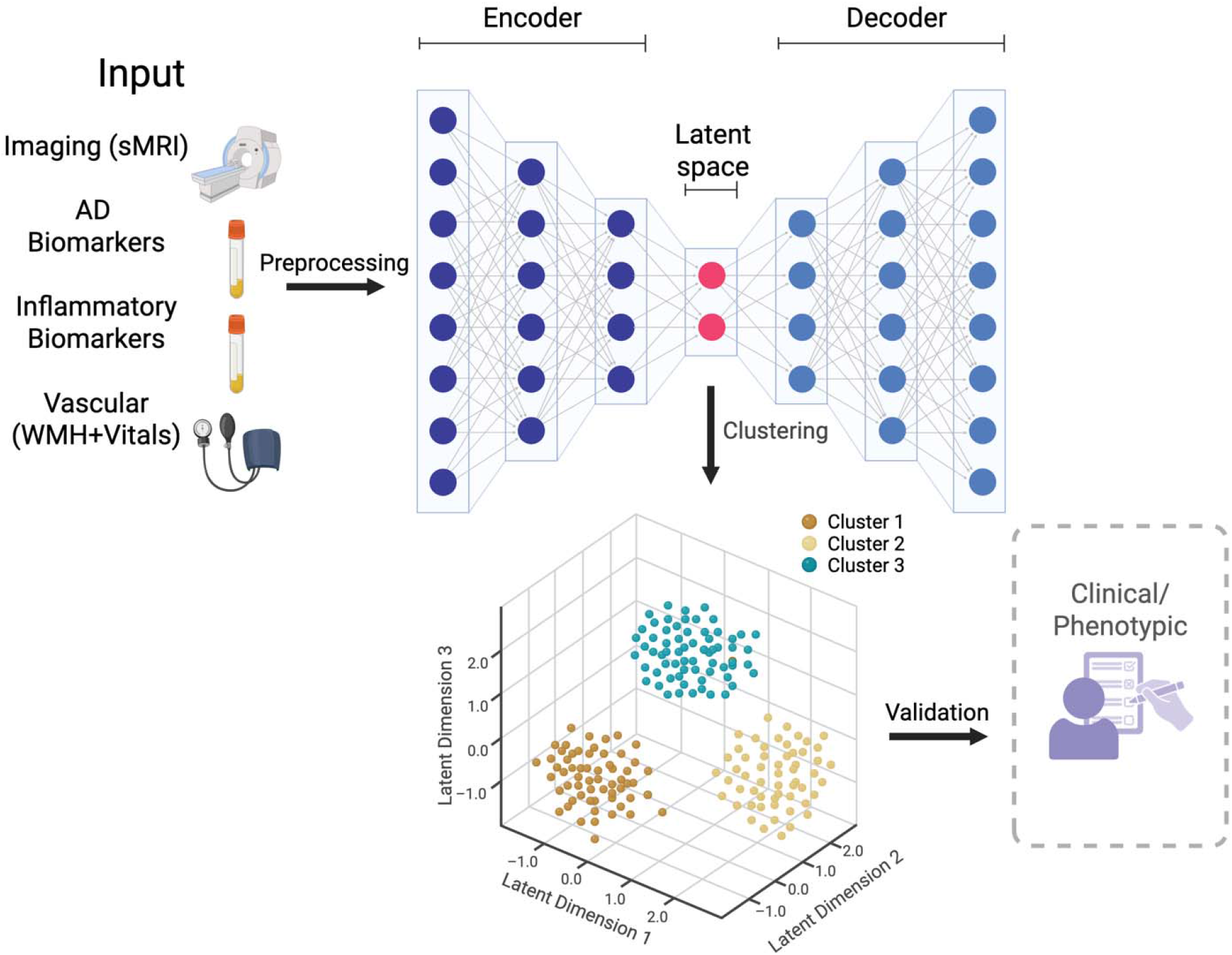
Study Outline. A modified Deep Embedded Clustering (DEC) approach was used to learn a compressed, non-linear latent representation of a set of multimodal neuroimaging (cortical thickness), plasma AD biomarkers, plasma inflammatory biomarkers and vascular measures. The latent space representations were used to cluster subjects in 3 groups (determined, along with the number of latent dimensions via a comprehensive pre-training search). Cluster assignments were then validated against cross-sectional and longitudinal phenotypic data, unseen by the modified DEC algorithm, including data from an external dataset.

## Results

We analyzed baseline cross-sectional multimodal neuroimaging, biomarker and phenotypic data from a sample of 1136 participants (ages 50 to 90, mean: 65.58 ± 8.24, 753 females) from the Health and Aging Brain Study-Health Disparities (HABS-HD) dataset.

## Deep Embedded Clustering of Multimodal Data from Cognitively Normal Adults

A DEC approach was first applied to compress 52 features (selected from the original 79 features, using category-wide PCA-based feature selection, see Methods) into 4 latent dimensions and then cluster the entire sample into 3 groups (**Fig. 2A**). The number of clusters (K) and latent dimensions(d) were determined through a systematic pre-training search procedure (**Supplementary Fig. 1**). Cluster assignments were highly stable over retraining runs (**Supplementary Fig. 2**), with mean Adjusted Rand Index (ARI) of 0.972 obtained for pairwise partition comparisons (1225 pairs), and 0.969 for agreement with the primary partition used throughout the study.

**Figure 2.**
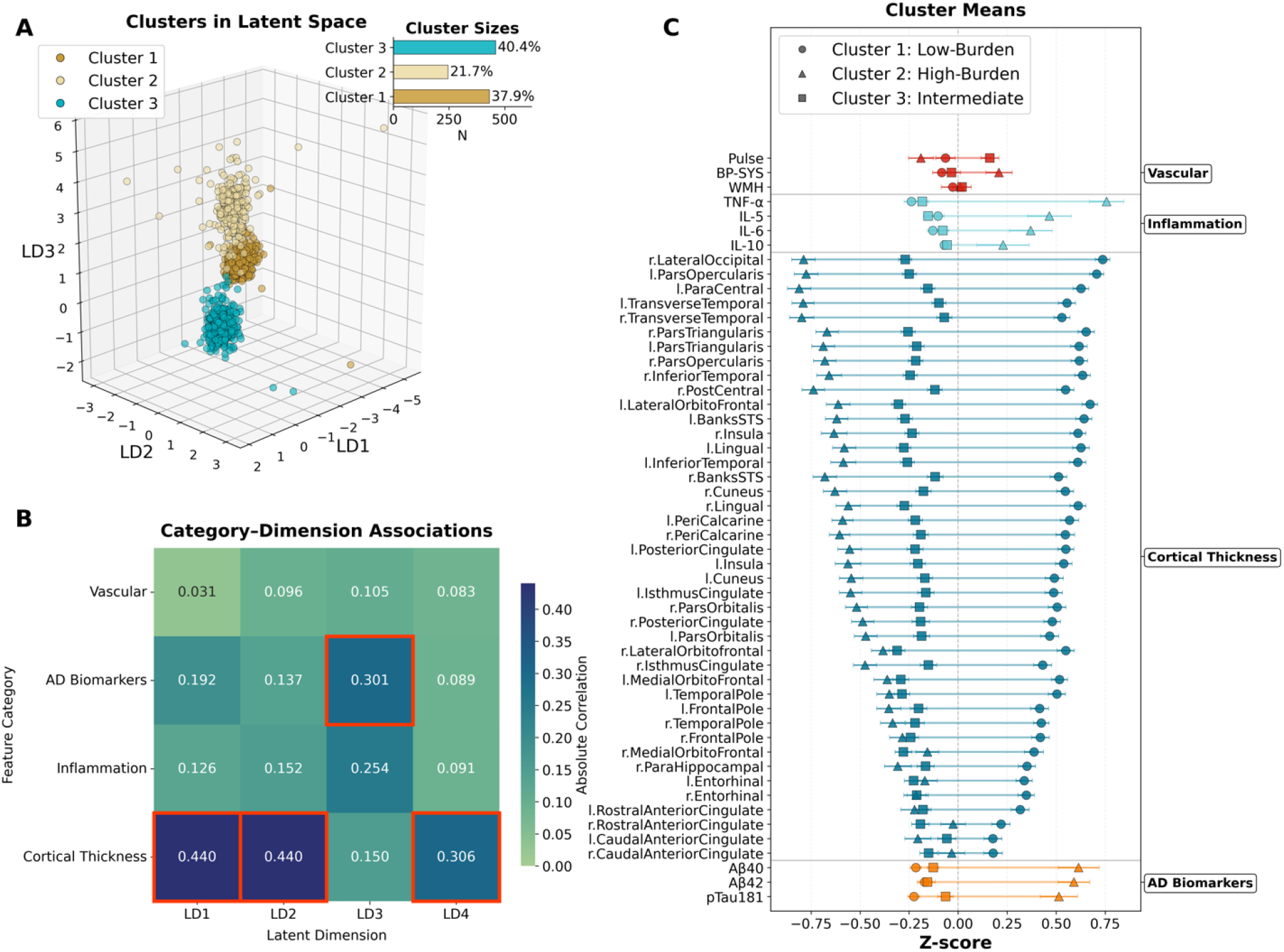
Clusters resulting from the modified DEC pipeline. (A). A total of 1136 subjects with available data were clustered into 3 groups, shown here on top of the first 3 latent dimensions. (B). The 4 feature categories loaded differentially onto the 4 latent dimensions learned by the model. Cortical thickness features loaded most strongly onto 3 of the latent dimensions while AD biomarkers loaded most strongly onto one latent dimension. (C). Plotting individual feature loadings for each of the 3 clusters reveals stratification along a gradient of biomarker burden (low, intermediate, and high).

Cortical thickness features loaded most strongly onto 3 of the 4 latent dimensions (**Fig. 2B**), while AD biomarkers loaded most strongly onto one latent dimension (**Fig. 2B**). Considering basic demographic attributes, subjects in the 3 clusters differed in age (F_2, 1133_ = 117.632, p < 0.001) and gender (χ^2^_2_ = 35.541, p < 0.001), with Cluster 2 having older subjects and a more balanced male/female ratio (1/1.02, compared to 1/2.48 and 1/2.35 in Clusters 1 and 3). The 3 clusters also differed in ethnoracial background (χ^2^_2_ = 15.410, p = 0.0039), with black participants overrepresented in Cluster 1 and underrepresented in Cluster 2. No differences were observed in education levels (F_2, 1133_ = 2.631, p =0.072).

Across all feature categories, Cluster 2 showed the highest burden in 46 of 52 features (88.5%). That is, subjects in this cluster showed higher levels in most inflammatory, AD and vascular features, and lower levels in cortical thickness features, indicative of cortical thinning. Mirroring these results, Cluster 1 showed the lowest burden in 50 of 52 features (96.2%). Cluster 3 was generally between these burden levels, although when considering single feature categories, the distinction between clusters 1 and 3 was not uniform. Taken together, the data indicate a stratification of clusters along a gradient of biomarker burden (low, intermediate, and high).

## Feature- and Feature Category-Level Contributions to Cluster Membership

Our clustering approach relied on features from multiple modalities/categories. We next sought to determine the extent to which individual features and the different feature categories contributed to the model’s clustering output. To that end, we trained a Random Forest classifier to predict cluster membership from all input features and applied SHapley Additive exPlanations (SHAP)^29^ to quantify interpretable, feature-level contributions to the clusters identified by the DEC approach. When averaging SHAP values across cluster assignments, cortical thickness, inflammatory (TNF-α) and AD biomarkers (Aβ_40_ and Aβ_42_) emerged as strong contributors to cluster membership, with cortical thickness features in frontal, insular, and inferior and medial temporal cortices dominating the top-ranked features (**Fig. 3A**). When considering SHAP value in single clusters, Cluster 2 (High-Burden) relied on a diverse set of features from all 3 of the 4 feature categories, while Clusters 1 and 3 relied on a more restricted set of features (**Supplementary Fig. 3**). Next, we assessed the contribution of the different feature categories to clustering assignments. SHAP values were then averaged for each feature category within each of the 3 clusters (**Fig. 3B**). Cortical thickness features showed the strongest contribution to cluster assignments, followed by inflammation features. Interactions between feature categories (**Fig. 3C**) were generally small in magnitude, suggesting that feature categories contributed to cluster assignments mostly independently. The largest interactions were found between inflammation and cortical thickness and between AD biomarkers and cortical thickness, again pointing to the diversity of feature contributions to the formation of clusters reported here. Vascular features showed relatively weaker interactions with other feature categories.

**Figure 3.**
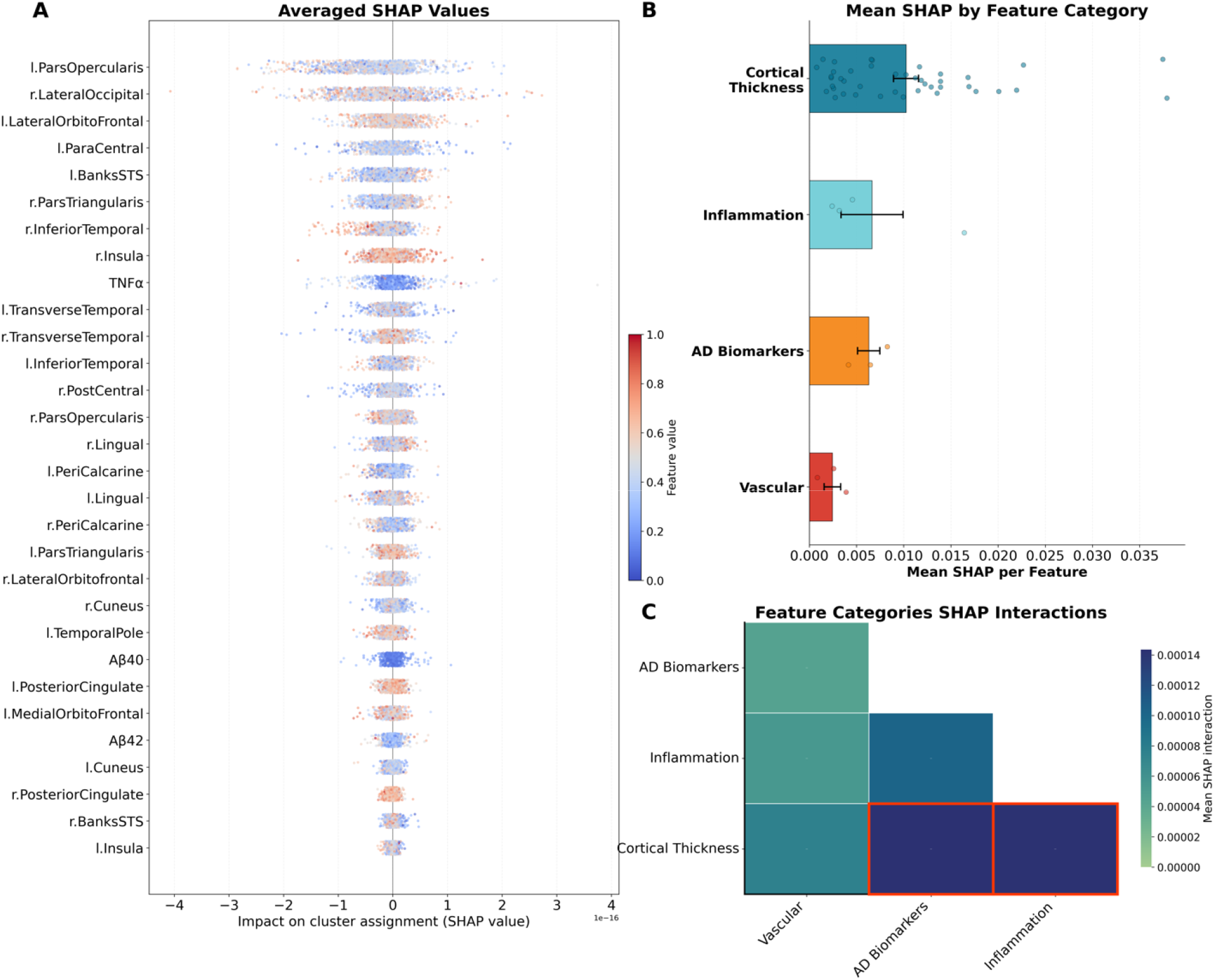
Feature- and Feature Category-Level Contributions to Cluster Membership. (A). The impact of individual features on cluster assignments is visualized by averaging SHAP values across cluster assignments. A mixture of cortical thickness, inflammatory and AD biomarkers contributed most strongly to cluster assignments. (B). SHAP values were averaged for each feature category, over each of the 3 clusters. (C). Mean absolute pairwise interaction values were averaged across all feature pairs, and clusters to assess for the contribution of feature category interactions to cluster assignments.

## Feature- and Feature Category-Level Contributions to Cluster Boundaries

We next determined the relationship between individual features and distances to cluster centroids, defined as the Euclidean distance in latent space between each subject and their assigned cluster’s center. We posit that this analysis can help elucidate the contributions of features and feature categories to cluster boundaries. When considering individual features, inflammatory biomarkers emerged as central contributors to cluster boundaries, with 4 of the 10 features showing the strongest associations with distance to cluster centroid being inflammatory biomarkers (IL-10, IL-5, IL-6 and TNF-α, **Supplementary Fig. 4A**). Similarly, when averaging distances across feature categories, the strongest associations with distance to cluster centroid were found for inflammatory biomarkers (**Supplementary Fig. 4B**), followed by vascular features.

## Phenotypic Comparisons of the Different Clusters

Data-driven clustering is inherently exploratory and may be unstable. We therefore next validated the DEC-derived clusters against phenotypic data unseen by the DEC pipeline. Given the widely documented role of cardiometabolic diseases in brain health and cognitive aging outcomes we first evaluated differences between clusters in the prevalence of cardiometabolic diseases^30,31^ (**Fig. 4A**). Differences between clusters were found in rates of diabetes (χ^2^_2_ = 18.215, p < 0.001), hypertension (χ^2^_2_ = 20.331, p < 0.001), and cardiovascular disease (CVD) (χ^2^_2_ = 23.120, p < 0.001) (**Fig. 4A**), with Cluster 2 (High-Burden) showing higher rates in all 3 diagnoses. In contrast, no differences were observed in rates of dyslipidemia (χ^2^_2_ = 1.608, p = 0.44). As cognitive decline in aging strongly associates with both depression^32,33^ and anxiety^34,35^ we also assessed whether the clusters differed in the rates of these two diagnoses (**Supplementary Fig. 5**). However, no significant differences were observed for either depression (χ^2^_2_ = 0.664, p = 0.72) or anxiety (χ^2^_2_ = 0.076, p = 0.96).

**Figure 4.**
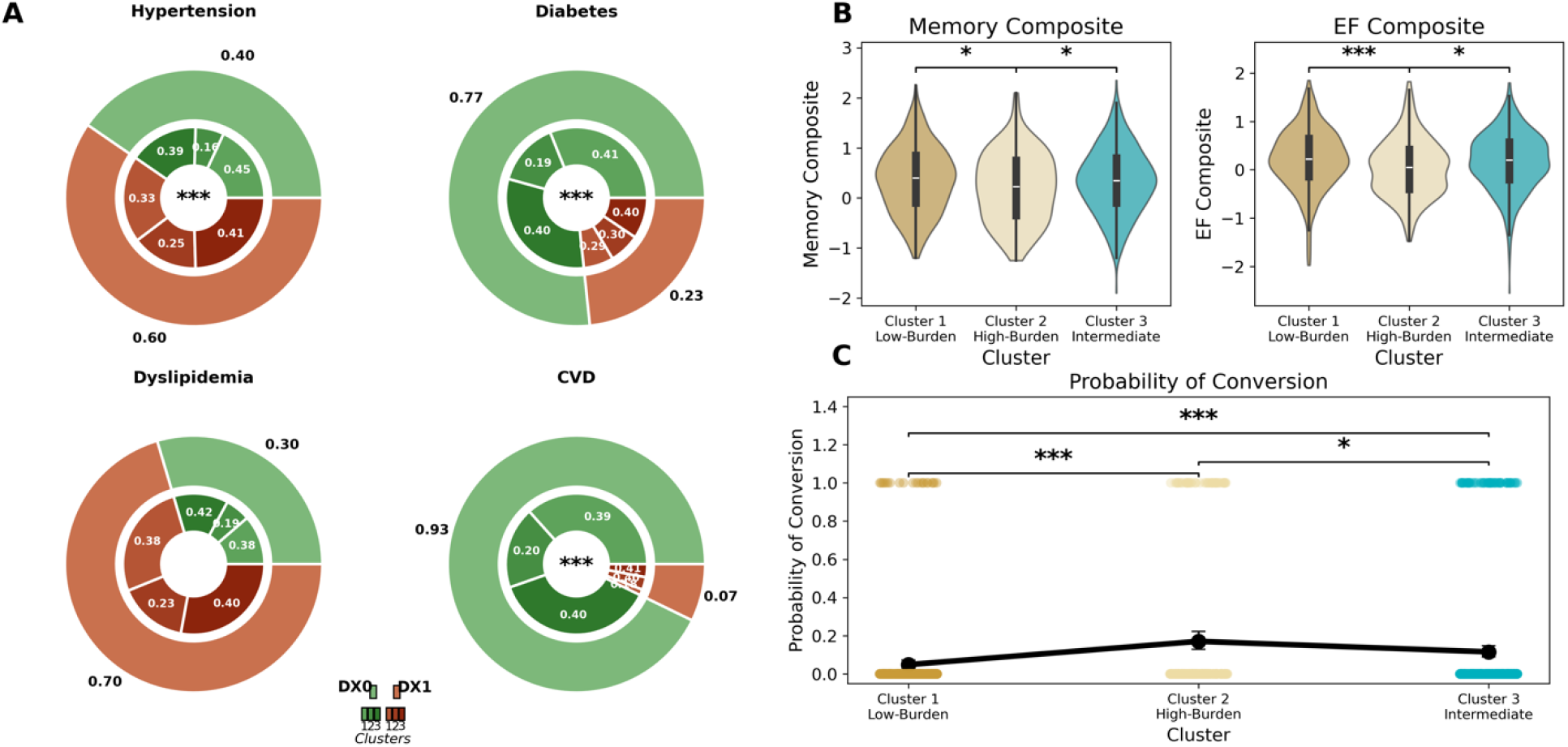
**Phenotypic Comparisons of the Different Clusters. (**A). Subjects assigned to the 3 clusters showed differential rates of hypertension, diabetes and cardiovascular disease (CVD), with Cluster 2 (High-Burden) showing the highest rates in each of the diagnoses. No differences were observed in rates of dyslipidemia. (B). Comparison of memory and executive functioning performance, measured via composite scores, revealed significant differences among the 3 clusters, with Cluster 2 (High-Burden) showing the overall worst performance. (C). Rates of longitudinal conversion from cognitively normal status to mild cognitive impairment or dementia differed between the 3 clusters, with Cluster 2 (High-Burden) showing the overall highest rates of conversion followed by Cluster 3 (Intermediate) and Cluster 1 (Low-Burden).

We next evaluated whether cluster membership was associated with differences in cognitive performance (**Fig. 4B**), using composite scores for memory and executive function as measures of cognition^36^. Significant differences were observed in memory performance (F_2, 1130_ = 4.495, p = 0.0114), with Cluster 2 showing significantly lower performance relative to both Cluster 1 (p= 0.011) and Cluster 3 (p=0.036). Since our focus was on age-associated cognitive decline, we additionally tested whether the observed group differences were not merely the result of age differences between the clusters. We found that the group differences in memory performance were retained (p=0.032) when controlling age. Significant differences were also found in executive functioning (F_2, 1131_ = 7.135, p < 0.001), with Cluster 2 showing significantly lower performance relative to both Cluster 1 (p=0.005) and Cluster 3 (p= 0.0169). The observed group differences were again retained when adjusting for age (p= 0.0101). Follow-up phenotypical data, acquired approximately 2 years post-baseline was then used to determine conversion from CN to MCI or dementia. Significant differences were observed between the 3 clusters in rates of transition along the cognitive decline spectrum (Likelihood Ratio Test:χ^2^_2_ = 27.889, p < 0.001), with subjects in Cluster 2 showing higher conversion rates relative to subjects in Cluster 1 (p < 0.001) and Cluster 3 (p = 0.041). Differences were also observed in progression rates between Clusters 1 and 3 (p < 0.001). Cluster membership was strongly predictive of conversion rates, also after adjusting for age (p < 0.001).

## External Validation

We next set out to validate the clustering results reported above against an external dataset. We identified a sample of 100 CN subjects (ages 55 to 86, mean: 71.5 ± 5.92, 56 females) from Alzheimer’s Disease Neuroimaging Initiative (ADNI) ^37^ who had available data from a similar feature set as that used in the main analysis (98.1% overlap in features, **Supplementary Fig. 6**). The ADNI cohort is a multicenter longitudinal study that collects multimodal neuroimaging, biomarker, genetic, and cognitive data to characterize the progression of Alzheimer’s disease and identify early biomarkers of cognitive decline^38^. Cluster assignments were tested via out-of-sample inference, that is, by applying the encoder and cluster centers learned in the original training procedures without retraining (see Methods). Subjects in the external sample were divided into 3 groups, with a distribution that did not differ from the one observed in the main sample (χ^2^_2_ = 4.2996, p= 0.12; **Fig. 5A**). We subsequently determined rates of progression from CN to MCI and dementia, as observed over the course of all ADNI visits. Cluster membership significantly predicted progression rates (Likelihood Ratio Test: χ^2^_2_ = 14.499, p < 0.001, **Fig. 5B**), with Cluster 2 showing significant differences in rates of progression relative to Cluster 1 (p=0.001) and Cluster 3 (p =0.012). The significant effect of cluster membership was retained after adjusting for age (p < 0.001).

**Figure 5.**
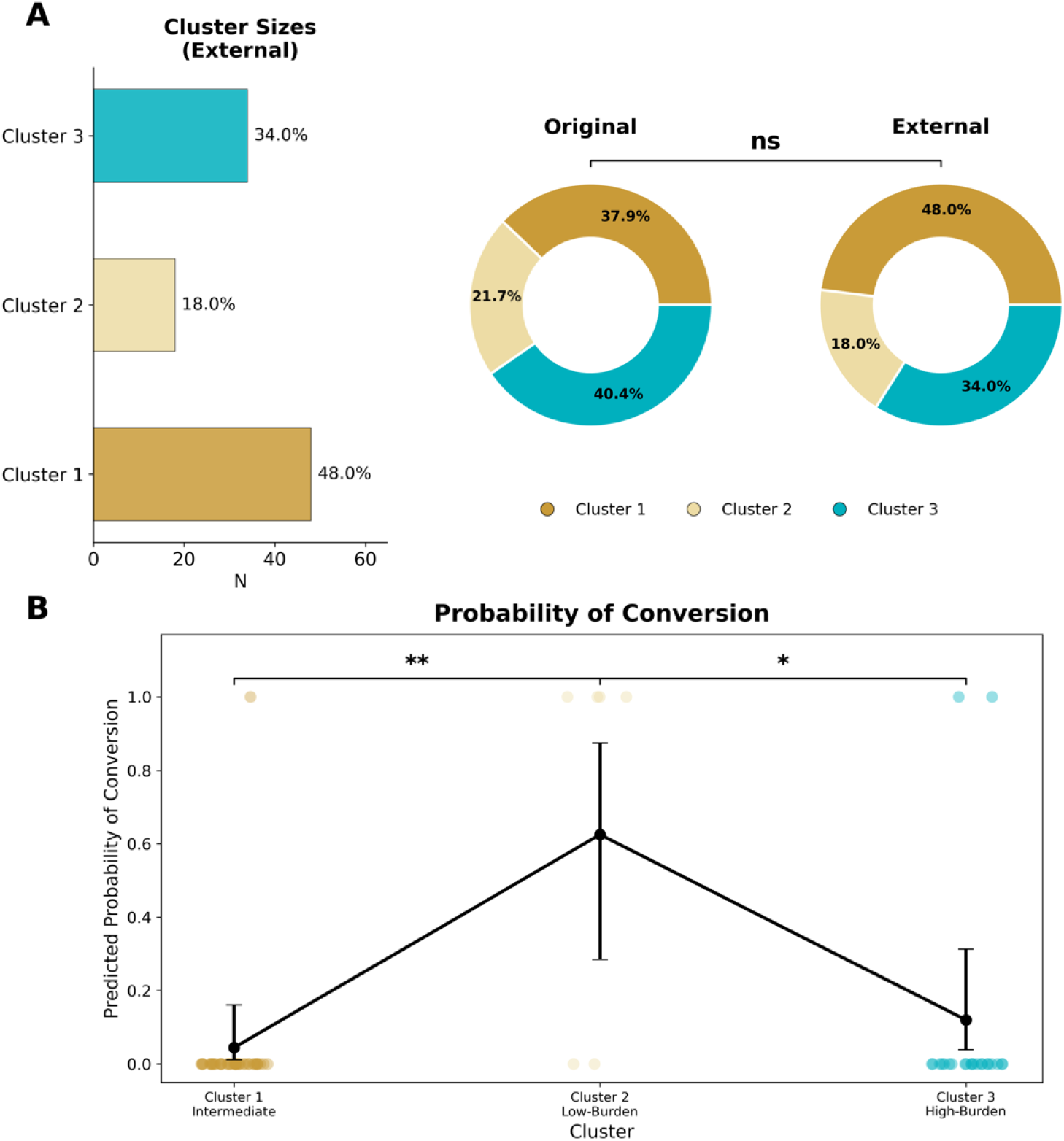
**External Validation of Clustering Results**. (A) Subjects in the external dataset (ADNI) were assigned to each of the 3 clusters via out-of-sample inference, wherein the encoder and cluster centers learned in the original training were applied directly without retraining. The proportion of subjects in each of the 3 clusters did not differ in the main and external datasets. (B). Conversion rates from cognitively normal status to mild cognitive impairment or dementia showed significant differences between the 3 clusters, with the highest conversion rates observed in Cluster 2.

## Discussion

In this study we used a DEC approach with multimodal data that identified three biologically distinct subgroups of cognitively normal adults. Our findings indicate that normal aging is not biologically homogeneous but is comprised of three distinct subgroups that represent a spectrum of burden across multiple neurobiological mechanisms, driven primarily by inflammation and cortical thinning^39–41^. The model-based subgroups were strongly differentiated along both cross-sectional cognitive performance (despite all being classified as cognitively normal) and conversion to MCI or dementia within two years.

Our findings highlight that individuals who are classified as “cognitively normal” may be on varying biological aging trajectories^42^. In particular, the DEC approach clustered participants along a gradient of biomarker burden, including: (1) a Low-Burden healthy aging subgroup (minimal inflammation, preserved cortical thickness, low AD pathology); (2) an intermediate vulnerability group (moderate inflammation and AD pathology, moderate cardiometabolic disease); (3) a High- Burden group likely at a preclinical phase for neurodegeneration (elevated burden of inflammation, cortical thinning, cardiometabolic disease). Critically, the model-based subgroups were shown to vary in clinically meaningful ways, with the High-Burden cluster showing the highest cardiometabolic disease burden, worse memory and executive function and the highest risk of progressing to MCI or dementia. The intermediate cluster, on the other hand, showed more preserved cognitive performance and favorable rates of cognitive decline, despite showing intermediate levels of biomarker burden. This may potentially reflect the operation of reserve mechanisms^43–45^, that allow the brain to withstand these levels of pathological burden.

These possibilities need to be examined in future research. Altogether, these results support the concept of a preclinical vulnerability state in which measurable neurobiological pathology is present before cognitive impairment is severe enough to be clinically detectable on neuropsychological testing.

We identified that inflammation appears to be a key factor that contributed to cluster formation and cluster boundaries, with TNF-α being the predominant cytokine that contributed to clustering, along with additional contributions of IL-6, IL-5 and IL-10 to cluster boundaries. Inflammatory biomarkers showed the strongest associations with distance from cluster centroid, which reflects their contribution to the biological boundaries of the cluster, and may better capture individual variation in disease states than discrete clustering assignments^46^. In prior studies elevated inflammatory cytokines, particularly IL-6, predicted subsequent cognitive decline^47,48^. Inflammatory biomarkers, such as IL-6 have also been shown to correlate with cortical thinning^49,50^, consistent with the reported interactions between these two feature categories reported here. Other pro-inflammatory markers, such as TNF-α are associated with worse cognitive performance among older adults^51^. Our findings and those of others support the hypothesis that inflammation may contribute to brain aging and exacerbate cognitive vulnerability as people age^52^.

Our analyses also suggest that cortical thickness, particularly in frontal, insular, and inferior and medial temporal cortices strongly contributed to clustering. These findings are consistent with previous reports on greater cortical thinning in cognitively normal adults that can precede the onset of cognitive decline^53^, and can predict future cognitive decline prior to clinical symptom onset^54^. Even among cognitively normal individuals, subtle cortical thinning is detectable and contributes to the cumulative risk profile of an individual^55^. Our findings highlight that structural brain changes, clustered together with other neurobiological changes, may precede the onset of clinically detectable cognitive impairment and may be an early marker of neurodegeneration.

We report differences in rates of cardiometabolic disease between the different clusters, such that the High-Burden cluster showed higher rates of hypertension, diabetes and cardiovascular disease. Cardiometabolic risk factors, including hypertension, diabetes, and cardiovascular disease, are associated with accelerated cognitive decline and increased incidence of dementia^56,57^. Similarly, midlife cardiovascular risk factors are associated with later-life brain atrophy and increased dementia risk^58^. We found no differences in dyslipidemia between clusters, which may indicate that not all cardiometabolic risk factors contribute to clustering, or that dyslipidemia is more heterogeneous and mitigated by treatment.

Cluster membership also differed by gender and ethnoracial background, with the High-Burden cluster showing a more balanced female-to-male distribution and a distinct ethnoracial composition relative to the low- and intermediate-burden clusters. These differences may represent differing biological, cardiometabolic, and social determinants of brain aging rather than intrinsic effects of demographic identity, consistent with previous findings that vascular and neurodegenerative biomarker associations vary across ethnoracial groups and gender ^59–61^.

One challenge in dementia prevention clinical trials is that cognitively normal older adults are highly heterogeneous, which can dilute the effects of an intervention if enrolled participants are at low short-term risk of cognitive decline (presumably members of the Low-Burden cluster in our study). Large multidomain prevention clinical trials, such as FINGER, MAPT and PreDIVA^62–64^, have demonstrated the difficulty in demonstrating benefit of these interventions across heterogeneous populations of varying cognitive risk^65,66^. Our approach suggests that multimodal biological clustering using an artificial intelligence approach may help identify a subgroup of cognitively normal adults on a high-risk cognitive decline trajectory who may be the most appropriate group to target for disease modifying or dementia prevention trials^67^.

The High-Burden cluster identified in our study exhibited both AD-related and structural brain changes, in addition to high inflammatory and cardiometabolic burden. The latter two factors may be potential biological targets for dementia prevention trials^68,69^ that may help mitigate conversion to MCI or dementia within two years. Prevention trials targeting vascular risk reduction (i.e. diet, exercise) or inflammatory pathways may benefit from selecting those cognitively normal individuals on the high-risk cognitive cluster. Our clustering approach may provide a framework for selectively targeting individuals using a biologically-informed risk stratification method in those classified as cognitively normal.

Several limitations of the current study are worth noting. First, the clustering assignments were obtained from cross-sectional baseline data, thus they likely represent biological profiles associated with different presumed levels of cognitive risk, which should still be validated against longitudinal biomarker data. Second, although external validation of our clustering pipeline using the ADNI cohort is a strength of the study, there are differences among participant demographics and recruitment structure between the HABS-HD and ADNI cohorts. Therefore, external validation in additional cohorts would be warranted in the future and could help identify whether cohort specific inclusion/exclusion criteria impact the results. Lastly, clustering is by definition an exploratory procedure, and future work would determine whether these clusters can be converted into clinical risk stratification tools in additional separate cohorts for external validation.

In conclusion, our study suggests that multimodal biological data in cognitively normal individuals can be clustered along a gradient of biomarker burden to detect subgroups with differential risk for future cognitive decline. Artificial intelligence-based methods can thus identify individuals with normal cognition who are on varying cognitive aging trajectories. Our findings may assist in detecting biological targets for cognitive decline and dementia, and most importantly provide an approach for identifying those cognitively normal individuals at highest risk of future cognitive decline who may benefit from enrollment in disease modifying or prevention trials.

## Materials and Methods

### Participants

Data from a total of 1136 participants were analyzed (mean age=65.58 ± 8.24, 753 females). Data were obtained from the Health and Aging Brain Study-Health Disparities (HABS-HD)^70^, an ongoing community-based prospective study aimed at understanding brain health and aging. The HABS-HD dataset is a longitudinal, community-based cohort designed to investigate biological and social determinants of cognitive aging and Alzheimer’s disease (AD) risk in a racially and ethnically diverse population. The study includes extensive multimodal phenotyping, including neuroimaging, blood-based biomarkers, cardiometabolic measures, and detailed cognitive assessments collected from non-Hispanic White, Black, and Mexican American participants, residing in the United States^71^. All participants were cognitively normal (CN) at baseline. Follow-up phenotypic data, acquired roughly 2 years post-baseline was used as well, to determine transitions to MCI and dementia.

Inclusion/exclusion criteria, detailed and discussed elsewhere^70^. In short, participants were age 40 and above, were able to provide informed consent, self-reported race/ethnicity of Hispanic, non-Hispanic White, or non-Hispanic Black, were willing to provide blood samples, willing and capable to undergo MRI and positron emission tomography (PET) scans and were fluent in English or Spanish. Exclusion criteria were Type 1 diabetes, current urinary tract infection (UTI) or uncontrolled inflammatory condition, diagnosis of cancer within the last 12 months, chemotherapy or radiation treatment within the same time period, active mental illness impacting cognition (excluding depression and anxiety), other serious medical condition impacting cognition, traumatic brain injury with loss of consciousness within the last 12 months, current alcohol or substance abuse and imaging contraindications. Additional multimodal data from a total of 100 participants (mean age=71.59±5.88), all cognitively normal at baseline, were taken from Alzheimer’s Disease Neuroimaging Initiative (ADNI). ADNI is a large, longitudinal, multicenter study designed to develop and validate clinical, imaging, genetic, and biochemical biomarkers for the early detection and tracking of Alzheimer’s disease to improve clinical trials^72^. All procedures were approved by the local Institutional Review Board, and all study participants provided written informed consent.

### Biofluids Data

Procedures for collection of fasting blood samples in the HABS-HD study are detailed elsewhere^70^. Samples were assayed using the Single Molecule Array (SIMOA) technology with Quanterix kits. Levels of Amyloid- β_40_(Aβ_40_), Aβ_42,_ and phosphorylated Tau_181_ (pTau_181_) were extracted and considered as biomarkers for AD pathology in the current study. Similarly, levels of IL-5, IL-6, IL-10, and Tumor Necrosis Factor-alpha (TNF-α) in plasma were extracted and included here as inflammatory biomarkers.

### Neuroimaging Data Acquisition and Analysis

Multimodal imaging data in HABS-HD were acquired with a 3T Siemens Vida or Skyra scanners. Here, we used: (a) Structural MRI data, acquired with a 3D T1-weighted magnetization prepared rapid acquisition gradient echo (MPRAGE) sequence (repetition time (TR) = 2300 ms, echo time (TE) = 2.93 ms, matrix = 256, field of view = 270, 1.2 mm slice thickness, voxel size = 1.1 mm × 1.1 mm × 1.2 mm). (b). T2-weighted fluid attenuated inversion recovery (FLAIR) sequence (repetition time (TR)= 4.800 ms, echo time (TE) = 441 ms, TI time = 1650 ms, matrix = 256, field of view = 256, slice thickness = 1.2 mm, voxel size = 1.0 mm × 1.0 mm × 1.2 mm). Cortical thickness measures were extracted from MPRAGE data using FreeSurfer ^73^ for 68 regions of interest from the Desikan-Killiany atlas^74^. The data are provided by HABS-HD as derived variables. Total volumes of WMH were quantified based on both sequences above using the lesion growth algorithm in the Lesion Segmentation Toolbox for the SPM software. Total WMH volumes, which were used as measure of vascular functioning (see below) were provided by HABS-HD as derived variables^75^.

### Vascular Measures

Four measures of vascular/cerebrovascular function were considered. Total volumes of WMH (see above), a proxy of small vessel disease^76^, were included, along with pulse, systolic blood pressure, and diastolic blood pressure.

Neuropsychological Test Data and Cognitive Diagnosis.

Neuropsychological test data were used to aid in the validation of the clusters identified by the modified DEC model. We considered separately measures of memory and executive function, following procedures used previously with HABS-HD data^77^. For memory performance, age and primary language-adjusted Z scores for the Logical Memory test in the Wechsler Memory Scale,3rd Edition (WMS-III): immediate and delayed recall, and the Spanish-English Verbal Learning Test (SVELT) were used. For executive function similarly adjusted Z score for the Trail Making Test Parts A & B total time, Letter fluency and Digit Span total score were used. These adjusted Z score were then averaged, resulting in composite scores for memory and executive function^77^. Consensus diagnoses of cognitive status were assigned to subjects based on self- or informant-based report of cognitive change, Clinical Dementia Rating (CDR) Sum of Boxes scores (CDR-SB=0 for CN, MCI=0.5–2.0, Dementia ≥ 2.5), and based on neuropsychological test results.

### Medical comorbidities

Comorbid medical conditions including hypertension, diabetes, CVD, dyslipidemia, depression and anxiety (along with several other conditions not analyzed here) were coded as present (1) or absent (0) based on (a) self-reported medical history, (b) clinical labs, (c) medication list, and (d) other objective measures via reviewed by a medical professional associated with HABS-HD

### Data Preprocessing

All features were first standardized using Z-score normalization. Features across all 4 feature categories were also subjected to PCA-based feature selection, with the objective of reducing the large imbalance in feature counts across categories, and minimizing redundancy within feature categories. This step, which was applied separately within each feature category, selected the minimum number of features whose linear combinations explained ≥ 90% of the category’s variance. This step reduces the total feature count from 79 to 52 (with cortical thickness features reduced from 68 to 42, vascular features reduced from 4 to 3, with no changes in the other feature categories).

### Deep Clustering Architecture

We employed a modified deep embedded clustering (DEC) approaches to learn the compressed non-linear latent representations of the multimodal set of features described above, and simultaneously cluster subjects into subgroups. This method first makes use of a deterministic autoencoder architecture to learn the low-dimensional latent representation of the data. The Encoder is comprised of an input layer, a first hidden layer with 64 units and Rectified Linear Unit (ReLU) activation, a second hidden layer with 32 units and ReLU activation, a latent layer of dimensionality *d* (determined pre-training, See below) with linear activation. The Decoder was comprised of a first hidden layer with 32 units and ReLU activation, a second hidden layer with 64 units and ReLU activation and an output layer with linear activation. Gaussian noise (σ = 0.05) was additionally added during training to prevent deterministic collapse and improve model robustness

Model training employed a composite loss function combining four objectives: (1) A *reconstruction loss* (mean squared error) aimed at preserving the correspondence between input features and autoencoder outputs; (2) A *clustering loss* that aimed to sharpen cluster separation (minimizing the KL divergence between soft cluster assignments Q and target distribution P, refined iteratively); (3) A *KL divergence regularization* which added a penalty for deviation of the latent representation distribution from a standard Gaussian prior N (0,1). This discouraged dimensional collapse and encouraged well-structured latent; (4) A *spectral regularization* term that penalized off-diagonal entries in the latent covariance matrix, and promoted orthogonal, disentangled feature representations. Loss weights used in the final version of the pipeline were set to λ_reconstruction_ = 1.0, λ_clustering_ = 1.0, λ_KL_ = 0.005, and λ_spectral_= 0.03. Our overall implementation modifies the original DEC framework^28^, adding KL and spectral regularization terms. This was motivated primarily by the multimodal, high dimensional structure of the input data used by the model for clustering (for related modifications to the DEC framework, see ^78^)

### Model Training and Clustering Procedures

The number of latent dimensions (*d*) and the number of clusters considered (*K*) were first selected via a systematic two-stage search. First, the number of latent dimensions was selected from a range of values (2-6) by evaluating the reconstruction error of the autoencoder, pre-training and using the elbow method to identify the optimal value. Second, the number of clusters was chosen from a range of values (2-5) by repeating the modified DEC pipeline (with the chosen number of latent dimensions from step 1), and choosing the optimal K value via silhouette scores, and the Calinski-Harabasz and Davies-Bouldin indices. Subsequently, training included two phases. First, the autoencoder was trained for 50 epochs with reconstruction loss using the Adam Optimizer (learning rate = 0.001, batch size = 32). Second, the model was then trained for 200 epochs with the full composite loss function. Clustering was first initiated using k-means on the initial latent representations, and subsequently updated every 3 epochs. Using a threshold silhouette of 0.2, ambiguous samples, if detected, were reassigned to the nearest cluster to reduce boundary ambiguity and improve cluster stability.

To assess the reproducibility of the DEC’s cluster solutions across multiple runs, the model was retrained for 50 iterations, and the Adjusted Rand Index (ARI) was computed as a measure of robustness, both pairwise, over 1,255 pairs, and when comparing each run to the primary partition used in the study.

### Features Contributing to Cluster Assignments (SHAP Values)

To identify which features, and feature categories contributed to discriminating between clusters a Random Forest classifier was trained to predict the cluster labels produced by the modified DEC pipeline, from all input features used by the pipeline. Five-fold stratified cross-validation was used to evaluate the quality of the classification performance, with balanced accuracy serving as the performance metric. SHAP values were then computed using the TreeExplainer method. Global feature importance was defined as the mean absolute SHAP value for a given feature, averaged across subjects and clusters. Importance scores were also averaged for feature categories. Interactions between feature categories were assessed by first computing pairwise feature interaction terms. The mean of absolute interaction values for all feature pairs from each of the possible category pairs were then averaged over subjects and clusters. The diagonal, denoting within-category self-interactions, was set to zero for visualization purposes.

### Implementation

Model training was performed in Python 3.11 using TensorFlow 2.21, scikit-learn 1.8 for clustering, and pandas 3.0 for data management.

### Statistical Analysis

Differences in continuous variables between clusters were evaluated with one-way ANOVAs, with significant effects followed by Tukey Honestly Significant Difference (HSD) post-hoc tests. Additional covariate-adjusted ANOVAs were conducted to adjust for relevant covariates, where necessary. Categorical demographic and clinical variables were compared between clusters using chi-squared tests of independence. Pearson’s correlation test was used to assess relationships between individual features and distance from cluster centroids. Finally, the predictive performance of cluster membership was examined using binary logistic regression, both unadjusted and adjusted for covariates, with model fit assessed via likelihood ratio tests. All statistical testing was carried out using Python 3.11 (statsmodels 0.14.6 and scipy 1.17)

### External validation

External validation of the clustering-derived cluster assignments was performed with data from ADNI. Data from all 4 feature categories used in the main modified DEC pipeline is provided by ADNI, with 98.1% overlap in features (**Fig. S6**). We first merged imaging, vascular, plasma AD biomarkers and plasma inflammatory biomarkers for subjects with a cognitively normal baseline diagnosis. Given high rates of missing data (primarily for plasma AD and inflammatory biomarkers which are available only for a subset of ADNI subjects), we systematically searched and retained for further analysis N=100 subjects with the least amount of missing data (ages 55 to 86, mean: 71.5 ± 5.92, 56 females). Prior to their inclusion in the external validation analysis, cortical thickness measure derived from ADNI were harmonized using the ComBAT method . Cluster assignments were applied to the external dataset via out-of-sample inference. Each sample in the external dataset was assigned to the nearest training cluster centroid by considering the Euclidean distance relative to the centroid in latent space. Label identities (Cluster 1, 2 and 3) were preserved in the external validation step to aid in interpretation. Chi-squared test compared the proportions of subjects assigned in the 3 clusters in the external dataset to those observed in the original training run. Differences between clusters in conversion rates to MCI or AD in the ADNI sample (stratified to resemble the lower conversion rates observed in HABS-HD) were again evaluated with binary logistic regression. Haldane– Anscombe correction was applied to prior to model fitting, since complete separation was observed (0 conversion in one cluster).

## Data sharing statement

All data are available through the HABS-HD (https://healthandagingbrainstudy.org/) and ADNI datasets (https://adni.loni.usc.edu).

## Supporting information

Diaz-2026-Supplemental

## Data Availability

All data produced are available online at:
1. https://healthandagingbrainstudy.org
2. https://adni.loni.usc.edu/data-samples/adni-data/

https://adni.loni.usc.edu/data-samples/adni-data/

## Acknowledgments

Research reported in this publication as part of the Health & Aging Brain Study: Health Disparities (HABS-HD) was supported by the National Institute on Aging of the National Institutes of Health under Award Numbers R01AG054073, R01AG058533, R01AG070862, P41EB015922 and U19AG078109. Data collection and sharing for this project was also funded by the Alzheimer’s Disease Neuroimaging Initiative (ADNI) (National Institutes of Health Grant U01 AG024904) and DOD ADNI (Department of Defense award number W81XWH-12-2-0012). ADNI is funded by the National Institute on Aging, the National Institute of Biomedical Imaging and Bioengineering, and through generous contributions from the following: AbbVie, Alzheimer’s Association; Alzheimer’s Drug Discovery Foundation; Araclon Biotech; BioClinica, Inc.; Biogen; Bristol-Myers Squibb Company; CereSpir, Inc.; Cogstate; Eisai Inc.; Elan Pharmaceuticals, Inc.; Eli Lilly and Company; EuroImmun; F. Hoffmann-La Roche Ltd. and its affiliated company Genentech, Inc.; Fujirebio; GE Healthcare; IXICO Ltd.; Janssen Alzheimer Immunotherapy Research & Development, LLC.; Johnson & Johnson Pharmaceutical Research & Development LLC.; Lumosity; Lundbeck; Merck & Co., Inc.; Meso Scale Diagnostics, LLC.; NeuroRx Research; Neurotrack Technologies; Novartis Pharmaceuticals Corporation; Pfizer Inc.; Piramal Imaging; Servier; Takeda Pharmaceutical Company; and Transition Therapeutics. The Canadian Institutes of Health Research is providing funds to support ADNI clinical sites in Canada. Private sector contributions are facilitated by the Foundation for the National Institutes of Health (www.fnih.org). The grantee organization is the Northern California Institute for Research and Education, and the study is coordinated by the Alzheimer’s Therapeutic Research Institute at the University of Southern California. ADNI data are disseminated by the Laboratory for Neuro Imaging at the University of Southern California. The content is solely the responsibility of the authors and does not necessarily represent the official views of the National Institutes of Health.

