## Supplementary material for "Deep multimodal clustering identifies biological subtypes of normal aging with divergent cognitive decline risk": Diaz-2026-Supplemental

**Supplementary Materials**

Monica M. Diaz^1^ and Eran Dayan^2^^; The Health and Aging Brain Study (HABS-HD) Study Team*, The Alzheimer’s Disease Neuroimaging Initiative

1. Department of Neurology, University of North Carolina at Chapel Hill, Chapel Hill, North Carolina, USA
2. Department of Radiology and Biomedical Research Imaging Center, University of North Carolina at Chapel Hill, Chapel Hill, North Carolina, USA

**^corresponding author:**

Eran Dayan, PhD:

**List of Supplementary Materials**

Figure S1 to Figure S7

**Supplementary Figures**

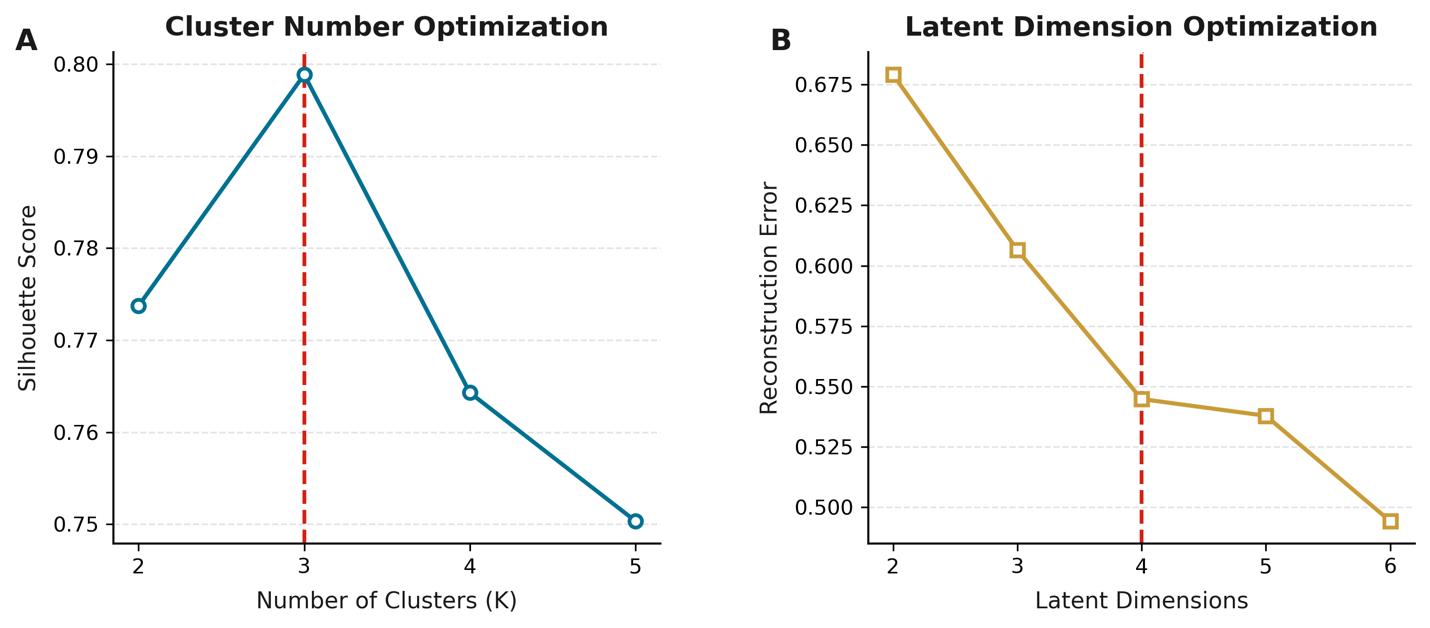

**Supplementary Figure 1. Optimization of hyperparameters used in the DEC pipeline.** (A). Silhouette scores as a function of cluster number K. (B). Reconstruction error as a function of the number of latent dimensions (2-6).

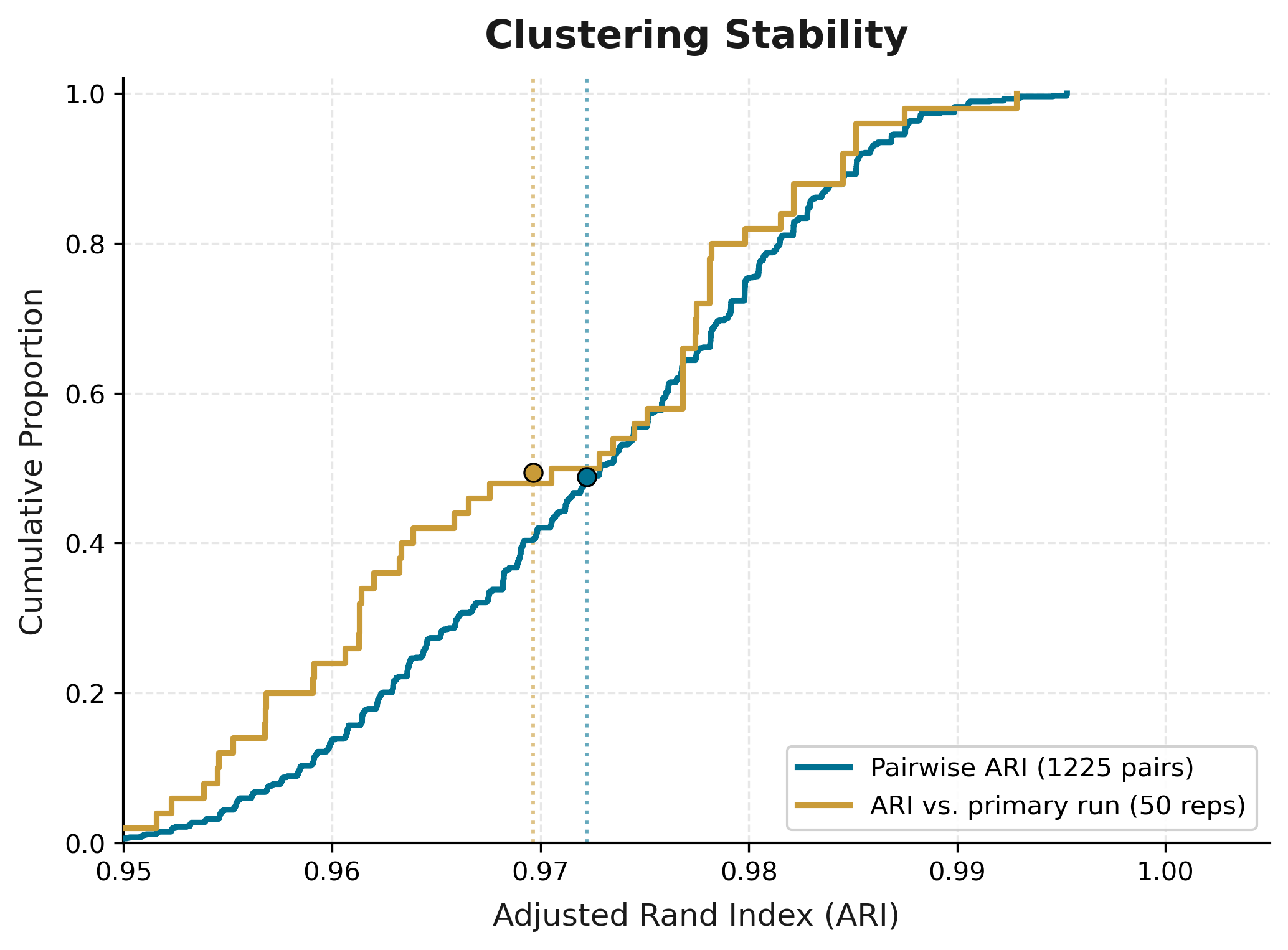

**Supplementary Figure 2. Clustering stability across multiple runs.** Adjusted Rand Index (ARI) values, obtained across 50 repetitions are shown, both as pairwise (1,225 pairs), and when comparing each run to the primary partition used in the study.

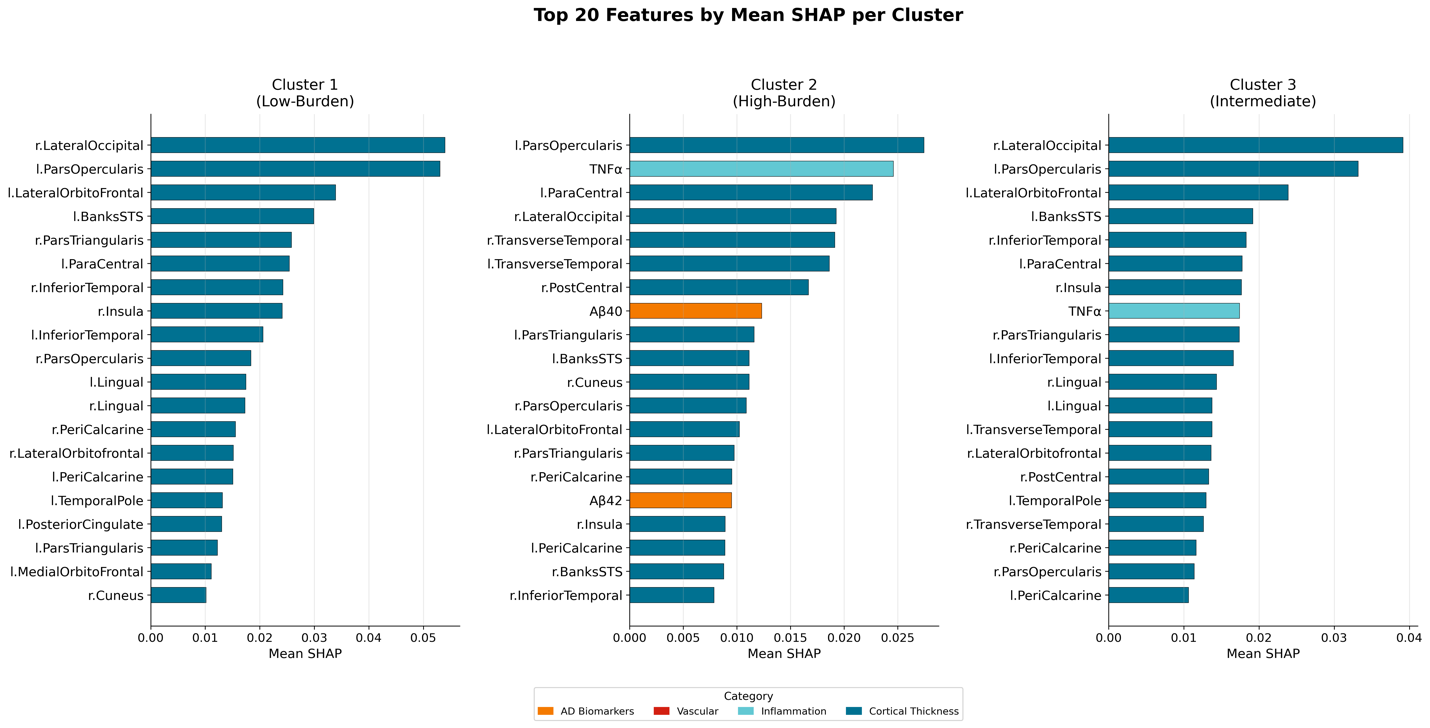

**Supplementary Figure 3. Feature-Level Contributions to Cluster Membership.** The impact of individual features on cluster assignments is visualized by displaying SHAP values (top 20) in each of the 3 clusters.

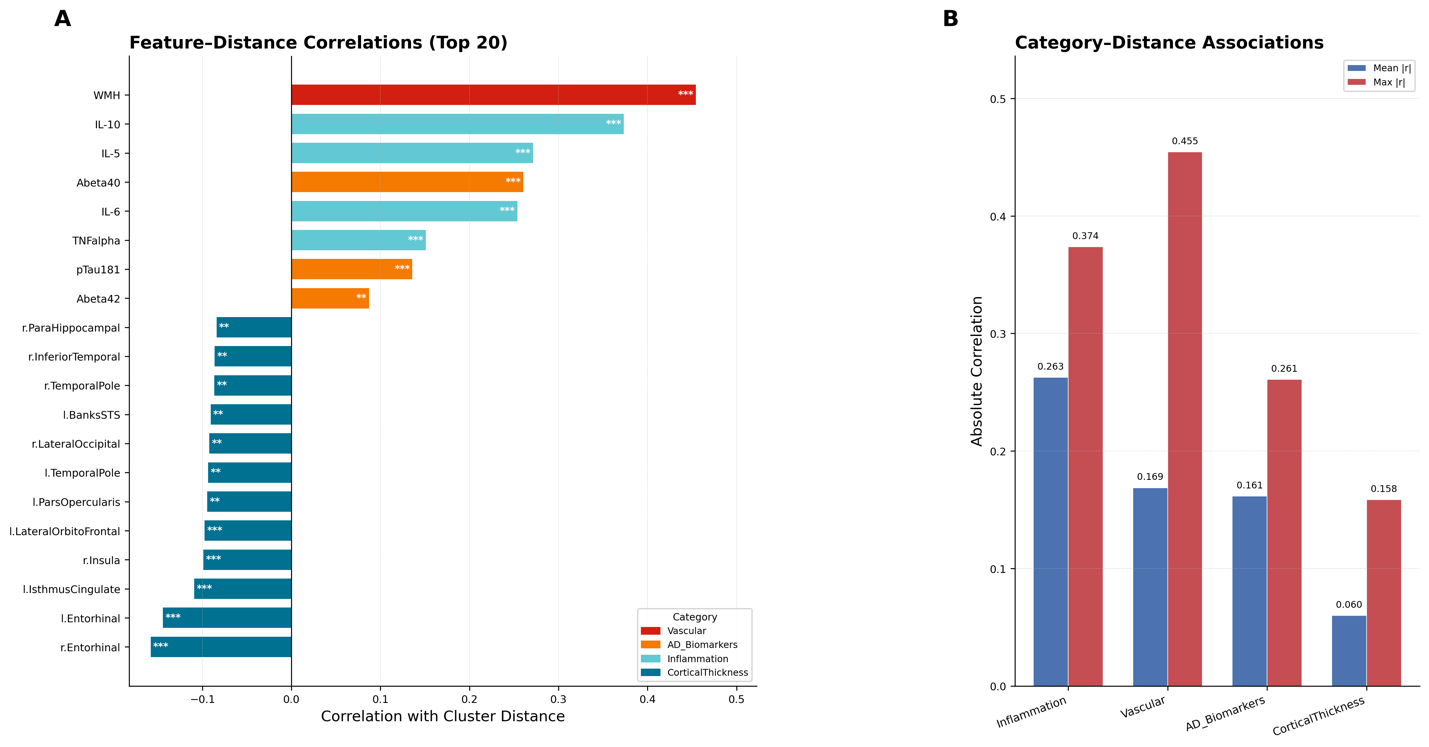

**Supplementary Figure 4. Feature- and Feature Category-Level Contributions to Cluster Boundaries.** (A). Correlations between single features and distance to cluster centroids. (B). Correlations between features and distance to cluster centroids, averaged over feature categories. Shown are both mean and maximal correlation strengths. * p < 0.05, ** p < 0.01, *** p < 0.001.

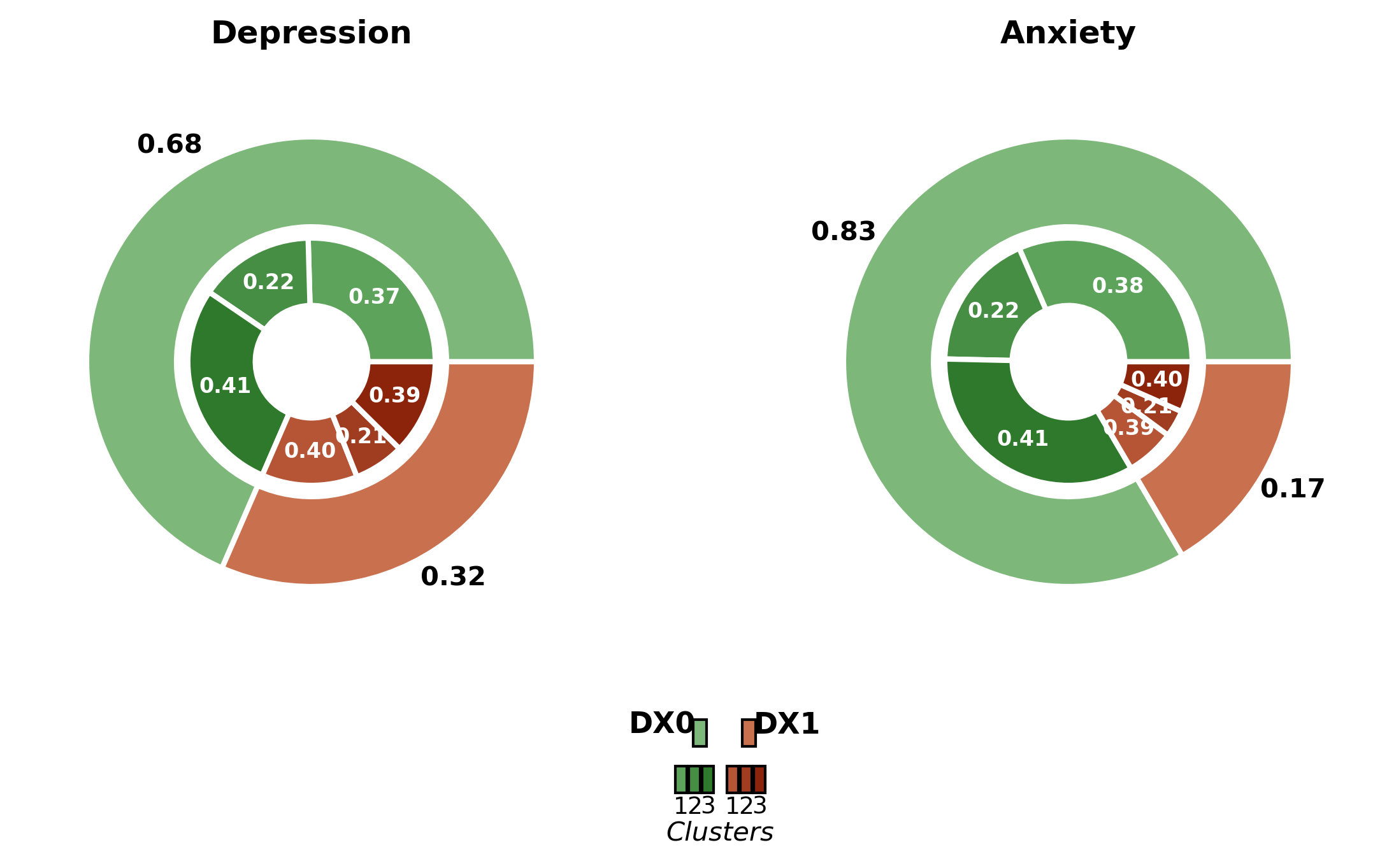

**Supplementary Figure 5.** Rates of depression and anxiety compared over the 3 clusters. Rates did not differ significantly for either diagnosis.

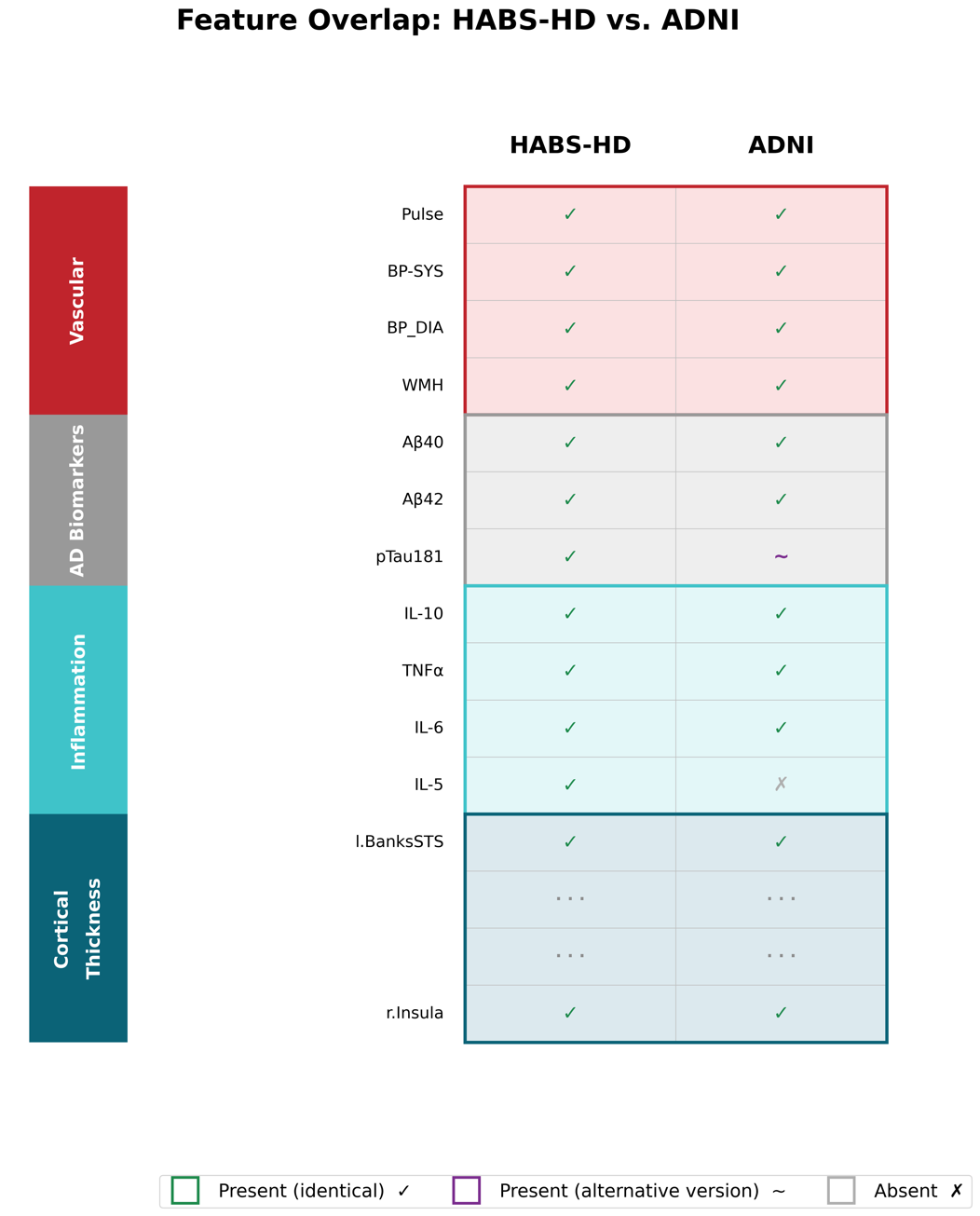

**Supplementary Figure 6. Comparison of features used in the HABS-HD and ADNI cohorts**. Features used in the main analysis (HABS-HD) and for external validation (ADNI) are shown for each of the 4 feature categories. Features are marked as present (identical in both cohorts), present with an alternative version (a single feature: plasma pTau181 in HABS-HD is replaced by plasma pTau217 in ADNI), or absent (available in HABS-HD and absent in ADNI).
